# Socio-economic status and outcomes after transcatheter aortic valve implantation: a systematic review and meta-analysis

**DOI:** 10.1101/2025.01.06.25320082

**Authors:** Sarah Verhemel, Majd Protty, Neil Ruparelia, Ahmed Hailan, Alexander Chase, Saud Khawaja, Nearchos Hadjiloizou, Iqbal Malik, Ghada Mikhail, Ramzi Khamis, Adam Hartley

**Affiliations:** National Heart and Lung Institute, Imperial College London, UK; Imperial College Healthcare NHS Trust, London, UK; Morriston University Hospital, Swansea, UK

**Author notes:** Corresponding author: Dr Adam Hartley, Vascular Sciences Section, National Heart and Lung Institute, Imperial College, Hammersmith Hospital, London, UK.

## Abstract

**Background:** Transcatheter aortic valve implantation (TAVI) procedures to treat severe aortic stenosis are rising, in line with aging populations and advancements in healthcare access. Economic development correlates with AS mortality rates. Socio-economic status (SES), defined by social and economic factors such as median household income, significantly influences cardiovascular outcomes. This study analyses the impact of SES on TAVI outcomes.

**Methods:** Following PRISMA guidelines, a comprehensive search was conducted across PubMed, Medline, Embase, Cochrane, ClinicalTrials.gov and Google Scholar, including literature up to August 1, 2024. The search employed keywords related to SES and TAVI. Of 393 identified studies, 127 were selected for full-text review, with ten addressing SES effects post-TAVI. Most studies had a retrospective design.

**Results:** The patient cohort comprised 319,557 individuals, (144,583 from low SES backgrounds and 174,974 from high SES backgrounds). The analysis revealed a higher burden of comorbidities in low SES patients. Post-TAVI, lower SES related to increased 30-day mortality, major adverse cardiovascular events and the need for pacemaker implantation, although it did not affect in-hospital mortality or 30-day readmission rates.

**Conclusion:** These findings underscore the substantial socio-economic disparities in TAVI outcomes and highlight the need for specific interventions to improve care for patients from less advantaged backgrounds.

**Key Learning Points:** *What is already known:*

- Lower SES negatively impacts cardiovascular outcomes due to disparities in healthcare access and comorbidities.
- TAVI is the standard treatment for severe aortic stenosis, especially in elderly patients.
- SES influences outcomes in cardiovascular procedures, but its impact on TAVI was unclear.

*What this study adds:*

- Low SES increases 30-day mortality, MACE, and pacemaker implantation rates after TAVI.
- SES does not affect in-hospital mortality or 30-day readmissions.
- Highlights the need for standardized SES metrics in TAVI research.

## Introduction

The overall prevalence of aortic stenosis (AS) varies widely, thought to be due to economic inequalities that result in limited healthcare access and reduced diagnosis rates [1, 2]. Severe AS is particularly common in individuals over 75 years of age, and this frequency is likely to rise due to aging populations, improvements in healthcare access and a resultant increased diagnosis [3]. Transcatheter aortic valve implantation (TAVI) has become the treatment of choice in this patient group due to reproducible outcomes, favourable valve durability data, patient choice and cost-effectiveness [3, 4]. Economic growth improves wealth indices and correlates with lower mortality rates from severe AS, largely due to better healthcare access, improved diagnosis, valve interventions, and a reduced prevalence of comorbidities driven by higher living standards [5]. Socio-economic status (SES), which combines various social and economic factors, is typically measured in the literature by categorizing populations based on median household income [6–8]. Inequalities in wealth distribution and low SES are known to exacerbate cardiovascular conditions and worsen outcomes [6–8]. However, the impact of socio-economic factors on outcomes following TAVI remains poorly understood.

Our aim is to review the effect of SES on outcomes following TAVI through a systematic review of the literature, supported by a meta-analysis of published studies.

## Methods

### Search Strategy

The study protocol was registered prospectively with the PROSPERO international registry (CRD42024588551). A comprehensive search of PubMed, Medline, Embase, Cochrane, Clinicaltrial.gov and Google Scholar was conducted in accordance with the PRISMA guidelines [9]. Relevant articles were reviewed up to August 1, 2024. Keywords and variations were used in medical subject headings (MeSH) or EMBASE (EMTREE) subject headings, including terms “socioeconomic status“[MeSH Terms], “socio-economic status”, “SES”, “social class”, “income”, “education level”, and “occupation”. These were combined with terms related to “transcatheter aortic valve implantation“[MeSH Terms], “transcatheter aortic valve replacement”, “TAVI”, “TAVR”, and “treatment outcome“[MeSH Terms], “outcomes”, “mortality”, “complications”, “readmission”, and “quality of life”.

### Study selection and eligibility

Two independent reviewers (SV, MP) assessed the relevance of studies by screening titles and abstracts. All study types relating to TAVI outcomes and SES were included, and the results were restricted to full-text articles only. EndNote™ was used as the reference manager, and full-text articles were manually screened. The co-primary endpoints were in-hospital and 30-day all-cause mortality. Secondary endpoints included major adverse cardiovascular events (MACE; as defined by each study’s specific definition), permanent pacemaker implantation (PPM), length of hospital stay, and 30-day readmission rate. The full search strategy is detailed in a flow diagram (Figure 1). Since there is no standardised measure for socioeconomic status (SES), we adopted each study’s specific definition, which encompassed factors such as education, literacy, housing, employment, insurance status, and neighbourhood median income.

**Figure 1:**
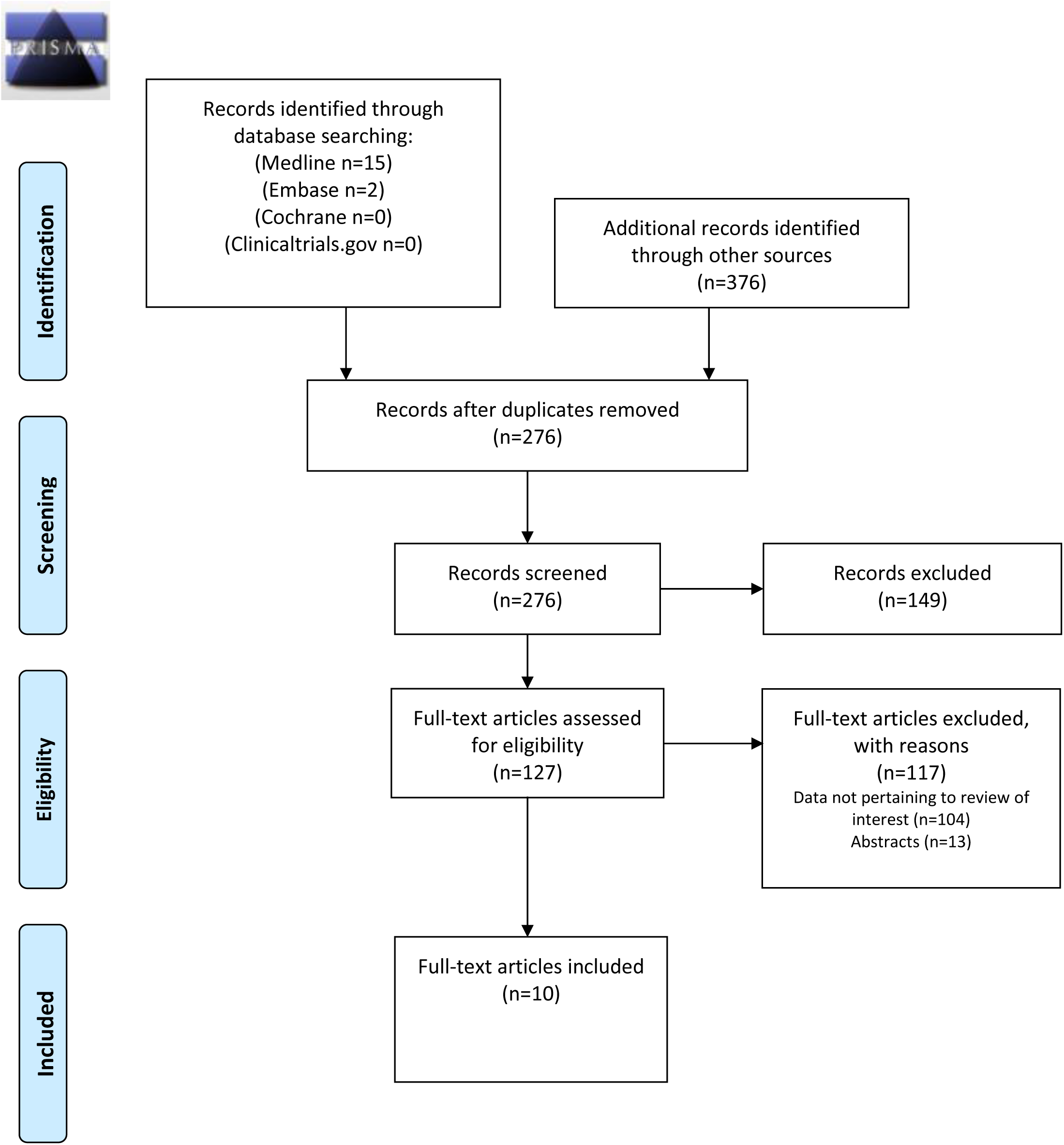
PRISMA Flow Diagram.

### Statistical meta-analysis

The characteristics of studies, comparison tables, and study data were stored and analysed using Revman 5.4.1™. Hazard ratios (HR) with 95% confidence intervals (CI) for all-cause mortality, MACE, and rehospitalization were calculated. Publication bias was assessed using ROBVIS™ incorporated in Revman™ desktop version, with unclear bias manually screened. A “traffic-light” table highlights the studies with low, unclear, or high risk of bias (Supplementary Table S1). Sensitivity analyses were conducted to investigate heterogeneity and the effect of individual studies on it.

## Results

A total of 393 studies included the relevant MeSH terms, of which 127 articles were selected for full text review. Only ten studies reported on socio-economic effects on outcomes following TAVI [10–19]. With the exception of the prospective study reported by Bilfinger et al [13], all other studies were retrospective regional population registries [10–12, 14–19]. Details on different SES grouping and studies included can be found in Supplementary Table S2. The vast majority of included studies had a low risk of bias (Supplementary Table S1).

### Pre-procedural Characteristics in Low vs. High Socio-economic Status

There were significant differences in the pre-procedural characteristics of patients undergoing TAVI across different SES groups. Among the studies, Hussain et al. [17] had the largest sample size of 296,740 participants, although specific pre-procedural characteristics were not provided. The average age of patients was slightly higher in the high SES groups [10, 11, 19]. In terms of sex distribution, the majority included in studies were males [10–14, 16–19]. However, Chabova et al. [15] reported 42% males in the low SES group and 60% in the high SES group. In general, patients from low SES backgrounds exhibited higher rates of comorbidity. Commonly reported conditions among low SES patients included hypertension, chronic obstructive pulmonary disease, diabetes mellitus and hyperlipidaemia. Additionally, these patients had higher prevalence rates of chronic kidney disease, coronary artery disease, arrhythmias and higher body mass index. Patient characteristics from the included studies can be found in Table 1.

**Table 1:** Summarized baseline characteristics of patients undergoing TAVI.

| Study (author, year, country) | Size (n) | Age, years mean ± SD/median, range | Low SES n (%) | Female n (%) | Diabetes n (%) | Hypertension n (%) | Atrial Fibrillation n (%) | BMI, kg/m<sup>2</sup> (mean ± SD) | Worse outcome Low SES | FU |
| --- | --- | --- | --- | --- | --- | --- | --- | --- | --- | --- |
| Czarnecki[14], 2017, Canada | 937 | 83 (78-87) | 18.2 | 44.3 | 48.5 | 95.6 | 31.9 | NA | No | 1 yr |
| Mohee[10], 2019, Wales, UK | 387 | 82.8 (8.3) | 23.5 | 45.2 | 25.7 | NA | NA | 27.3 (15.4) | No | 3 yr |
| Farwati[11], 2021, USA | 168853 | 81 (75-86) | 20.6 | 46.6 | 34.9 | 84.8 | 35.4 | NA | No | 30 D |
| Von Kappelgaard, 2021[12], Denmark | 13861 | N/A | 41.7 | 32.8 | N/A | N/A | N/A | N/A | Yes | 1 yr |
| Bilfinger[13], 2021, USA | 286 | 80 (9) | 25.5 | 48.3 | 39.2 | NA | 37 | 29.5 (5.8) | No | 3 yr |
| Chabova[15], 2022, CR | 439 | 77.4 (7.2) | 48.3 | 57.2 | 32.3 | 69.7 | 29.4 | 29.5 (5.7) | No | 2yr |
| Brlecic[19], 2023, USA | 142786 | 81 (75-86) | 19.4 | 48.5 | 41.3 | 89.8 | 55 | N/A | Yes | 1yr |
| Shawon[18], 2024, AUS | 4198 | 82.7 (7.5) | 15.3 | 40.6 | 33.8 | 88 | 55.9 | N/A | Yes | 1yr |
| Hussain[17], 2024, USA | 296740 | 78.7-79.9 | 21.1 | 44.5 | 37.7 | 89.7 | N/A | N/A | No | N/A |
| Li[16], 2024, USA | 27567 | N/A | 44.8 | 47.37 | 24.38 | 66.7 | 35.99 | N/A | Yes | N/A |

UK=United Kingdom, USA=United States of America, CR=Czech Republic, AUS=Australia N/A= not applicable, FU= follow up

## Outcome measures

### Socio-economic Differences and In-hospital All-cause Mortality

Six studies recorded in-hospital mortality post-TAVI divided according to SES groups [10, 11, 13, 15, 16, 19]. The total number of patients in the low SES group across all studies is N=75,316, with Brlecic et al. [19] having the highest rate of events, recording 410 events in the low SES group (N=27,791). The combined risk ratio (RR) for all studies is 0.94 (95% CI: 0.88, 1.01), with a Z-score of 1.74 and a p-value of 0.08, indicating no significant association between in-hospital all-cause mortality and SES (Figure 2A).

**Figure 2A:**
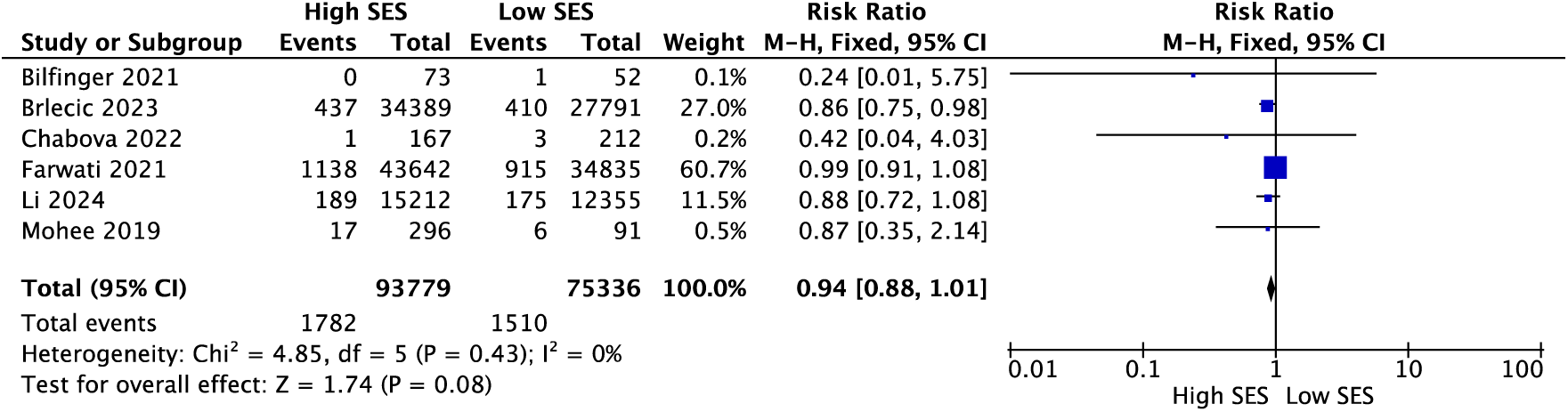
Forest plot of all-cause mortality in-hospital demonstrating no association between mortality and SES.

### Socio-economic Differences and 30-Day All-cause Mortality

Four studies included all-cause mortality during 30-day follow-up [10, 12, 13, 15]. The total number of patients in the low SES group across all studies is 1,420, with Mohee et al [10] having the highest rate of events, recording 88 events in the low SES group. The combined risk ratio (RR) for all studies is 0.77 (95% CI: 0.68, 0.87), with a Z-score of 4.12 and a p-value of <0.0001, indicating a significant association between increased mortality during 30-day follow up and low SES (Figure 2B).

**Figure 2B:**
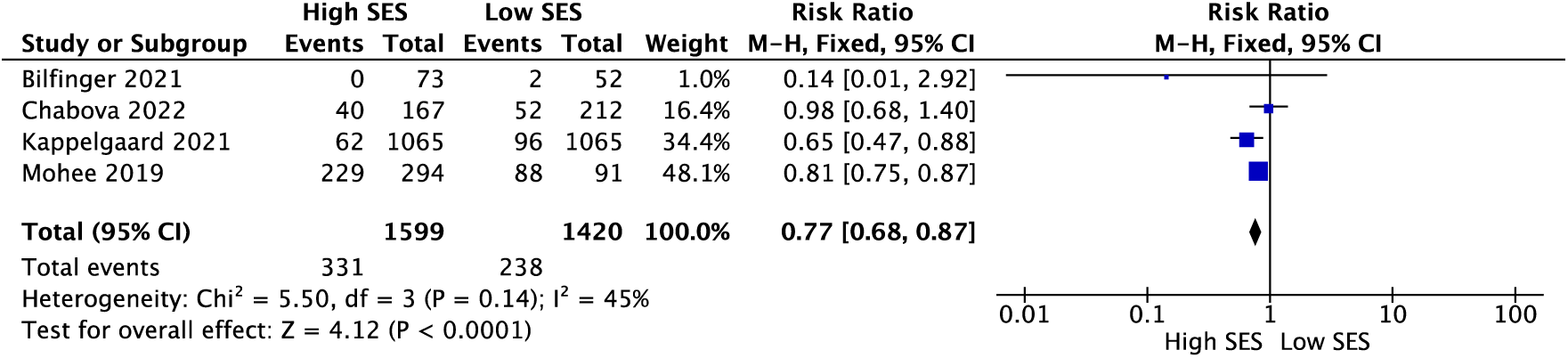
Forest plot of all-cause mortality at 30-day follow-up demonstrating an association between reduced mortality and high SES.

### Socio-economic Differences and Major Adverse Cardiovascular Events

Five studies recorded peri-procedural MACE divided according to SES groups [10, 11, 13, 15, 16]. The total number of patients in the low SES group across all studies is 47,621, with Farwati et al [11] reporting the highest number of events; documenting 6,045 events in the low SES group, accounting for 17% of their cohort of low SES patients. The combined risk ratio (RR) for all studies is 1.13 (95% CI: 1.10, 1.17), with a Z-score of 8.49 and a p-value of <0.00001, indicating a statistically significant association between increased peri-procedural MACE and low SES (Figure 3).

**Figure 3:**
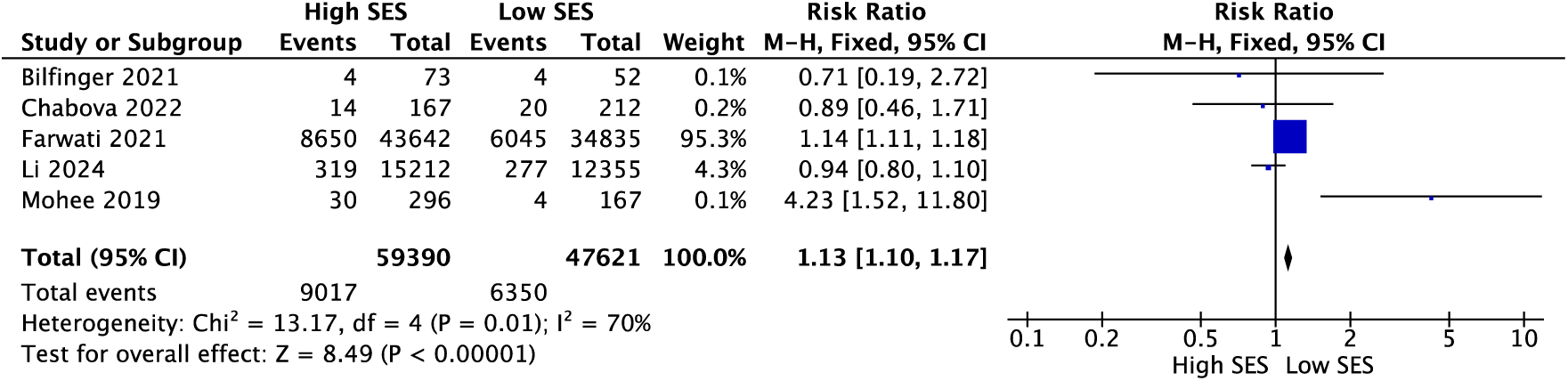
Forest plot of peri-procedural major adverse cardiovascular events demonstrating an association between high major adverse cardiovascular events and low SES.

### Socio-economic Differences and Peri-procedural Pacemaker Implantation

Five studies examined peri-procedural PPM insertion according to different SES groups [11, 13, 15–17]. The total number of patients in the low SES group across all studies is 110,278. Farwati et al [11] reported the highest number of peri-procedural PPM implantations in the low SES group, with 3,256 events out of 34,835 patients (9.3%). However, Chabova et al. [15] documented the highest percentage of peri-procedural PPM implantations, with 16.5% of their low SES cohort undergoing the procedure. The combined risk ratio (RR) for all studies is 1.21 (95% CI: 1.17, 1.26), with a Z-score of 10.08 and a p-value of <0.00001, indicating a statistically significant association between higher peri-procedural PPM insertion and low SES (Figure 4).

**Figure 4:**
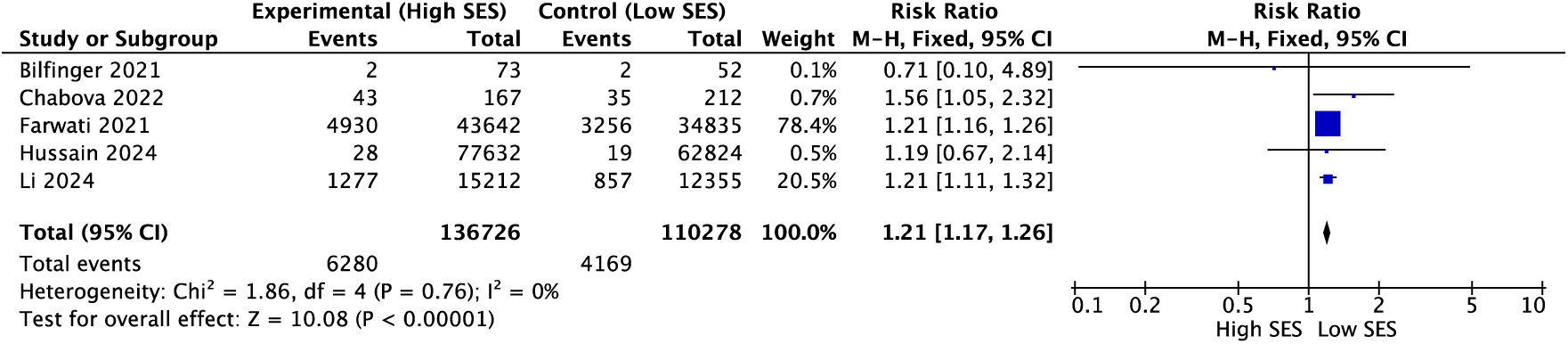
Forest plot of peri-procedural permanent pacemaker implantation demonstrating an association between high peri-procedural pacemaker insertion and low SES.

### Socio-economic Differences and Length of Hospital Stay

Five studies looked at total in hospital stay divided according to SES groups [10, 13, 15, 16, 19]. The total number of patients in the low SES group across all studies is 37,670, with Brlecic et al. [19] having the largest weight in the analysis. The longest length of in-hospital stay was reported by Mohee et al. [10] with an estimate of 29.7 days in low SES study population vs. 21.3 days in high SES group. The combined standardized mean difference for all studies is −0.23 (95% CI: −0.24, −0.21), with a Z-score of 32.30 and a p-value of <0.00001, indicating a statistically significant association between shorter peri-procedural hospital stay and higher SES (Figure 5).

**Figure 5:**
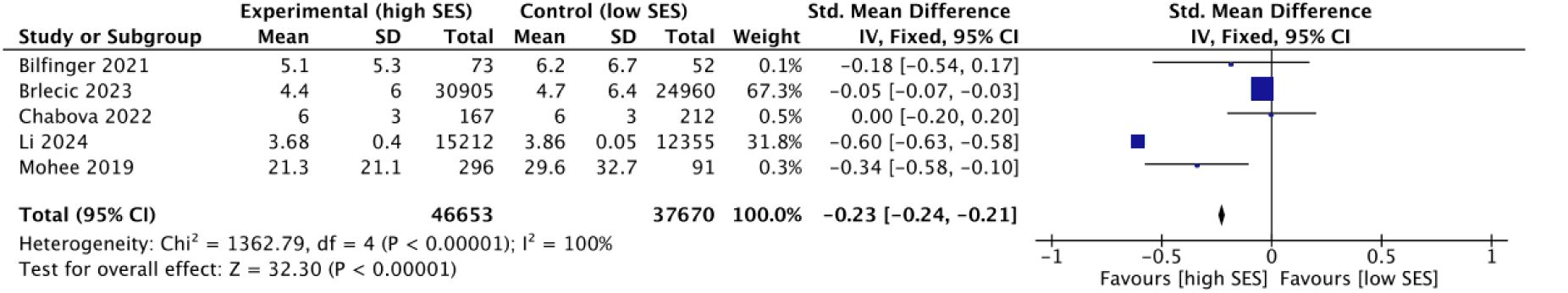
Forest plot of peri-procedural hospital stay demonstrating an association between shorter stay and higher SES.

### Socio-economic Differences and 30-Day Readmission Rate

Five studies examined data regarding 30-day readmission rate following TAVI divided according to SES groups [11, 13, 14, 18, 19]. Across all studies, the total number of patients in the low SES group was 60,645. Shawon et al. [18] recorded the highest percentage of 30-day readmissions, with 24% of their low SES cohort experiencing readmission within this period. The combined risk ratio (RR) for all studies is 0.98 (95% CI: 0.95, 1.01), with a Z-score of 1.30 and a p-value of 0.19, indicating no statistically significant difference in the 30-day readmission rate between high and low SES groups (Figure 6).

**Figure 6:**
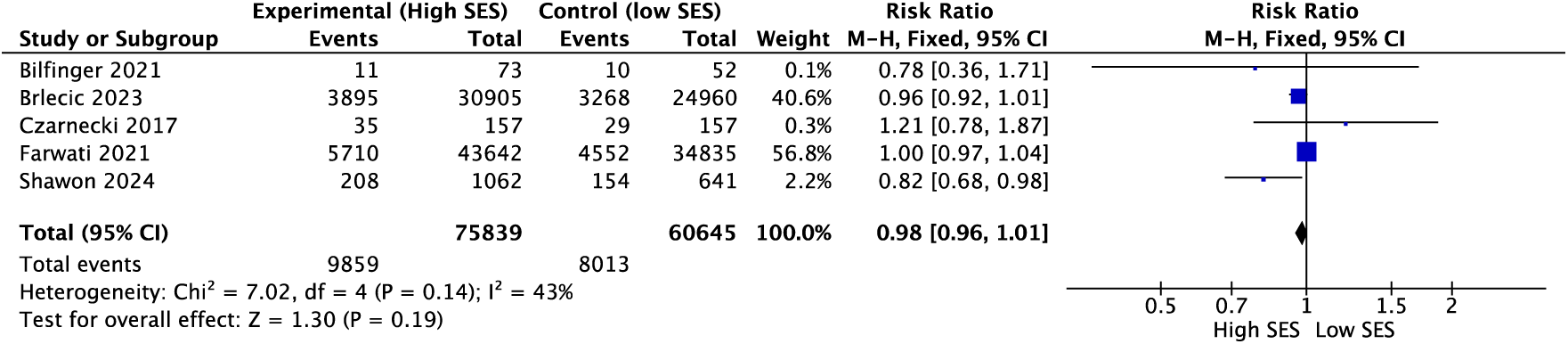
Forest plot of 30-day readmission rate demonstrating no association between readmission and SES class.

## Discussion

This meta-analysis evaluates the relationship between SES and outcomes following TAVI, particularly relevant given current global economic disparities and shifts [20]. Traditional TAVI research has often neglected to include SES as a baseline characteristic, which overlooks its potential impact on patient outcomes. Despite correlations established in prior studies between low SES and increased CVD risk [6, 8, 21], no standardized method for SES assessment in TAVI trials has been developed, hindering consistent correlation analysis.

The meta-analysis totals all available data and, contrary to individual studies that reported no significant SES-based disparities in hospital mortality, found a significant increase in 30-day all-cause mortality among patients with low SES [10, 12, 13, 15] (Figure 2). This aligns with findings from an isolated surgical aortic valve replacement (AVR) study [22] and research on area deprivation indices post-TAVI, which similarly identified neighbourhood conditions as critical predictors of mortality, as well as area deprivation index [23]. Unfortunately, the latter study was not included in the present analysis as it did not align with our study methods.

Our review highlights considerable variability across studies (Figures 2-6), likely attributable to differences in study design and SES definitions. This reinforces the necessity for standardized SES metrics to guide future research and individualized post-procedural care policies [24, 25].

The analysis shows that high SES groups have a lower risk of peri-procedural MACE, reduced 30-day mortality, and fewer PPM implantations compared to low SES groups. Although our data showed no significant difference in hospital readmission rates between the two groups, this lack of association between SES and readmission may be attributed to standardised healthcare protocols and policies aimed at reducing readmissions across all patient groups. It is notable, however, that the initial presentation appears to be divergent between SES groups. Studies by Chabova et al [15] and Li et al [16] reported significantly higher rates of urgent admissions for emergency TAVI in low SES populations, highlighting disparities in healthcare access and potential delays in symptom recognition. In contrast, Brlecic et al. [19] which includes the largest low SES cohort, found no significant association between SES and urgent TAVI admissions.

Conduction disturbances post TAVI are common complications with no clear consensus on predictive values [26, 27]. The higher incidence of permanent PPM implantation in low SES groups following TAVI may reflect regional variations in medical practice and clinical decision-making. Factors such as a higher prevalence of comorbidities in these populations and differing clinical thresholds for intervention could also explain this finding. Structural differences in healthcare access and policy also likely influence outcomes, warranting further investigation into these complex dynamics. Additionally, PPM insertion depends on various factors such as pre-existing bundle branch block, transcatheter heart valve design, and implantation depth, which are well-established predictors for PPM post-TAVI [27–31] but are not disclosed in most studies.

Healthcare costs increase significantly with prolonged hospitalisation, with a recent US study estimating an excess cost of 20 billion dollars due to extended hospital stays [32]. The association between low SES and prolonged hospitalisation highlights the economic burden faced by these patients. Longer hospital stays are strongly linked with lower SES, reflecting broader healthcare inequalities that extend beyond immediate medical needs. Factors often overlooked, such as the level of social support available at home, can delay discharge. For example, patients with limited family support or carers may need additional in-hospital care or longer recovery periods. Furthermore, a patient’s living environment is crucial; those lacking safe or suitable housing—such as flats or homes without stair lifts or with multiple flights of stairs—may encounter challenges to being discharged promptly. These socio-environmental factors underscore the need for a holistic approach to care planning and discharge protocols for patients from lower SES backgrounds to reduce disparities and improve outcomes. More prevalent comorbidities in lower SES groups may also contribute to prolonged hospital stays. Unfortunately, none of the reviewed studies provided reasons for extended hospitalisation or costs associated with it, likely due to their retrospective design.

## Socio-Economic Risk Framework

The extensive variability and the high degree of variance observed across the studies underscore the urgent need for standardised SES measures to better guide future research and inform targeted post-procedural care strategies [33]. We have proposed a framework that aims to effectively integrate SES considerations into risk stratification for TAVI outcomes (Figure 7). Building on the findings of Schultz et al. [34] regarding the impact of SES on cardiovascular health, our model incorporates key SES markers, such as income, education level, employment status, and neighbourhood deprivation. This SES risk framework system aims to complement existing clinical risk scores, offering a more nuanced and comprehensive assessment of patient outcomes once validated and adapted through future studies. The flowchart (Figure 7) illustrates how physicians could eventually incorporate these essential SES variables alongside established clinical risk scores, tailoring management to the SES needs of each individual, aiming to improve overall patient care and combat the SES inequalities demonstrated in this meta-analysis.

**Figure 7:**
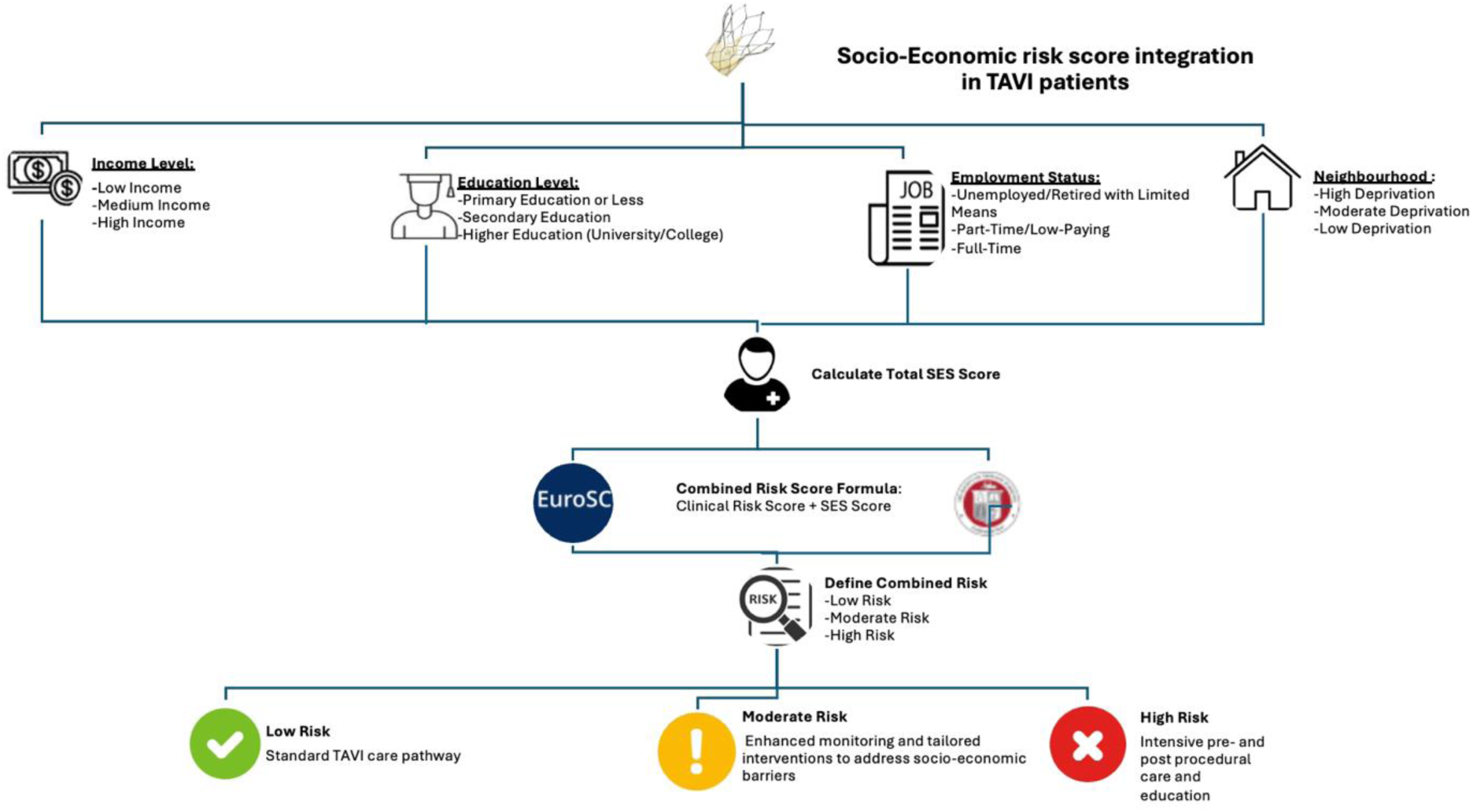
Flowchart allowing Socio-Economic risk Framework assessment in TAVI patients.

## Limitations

Studies included in this review share the same limitations. The key issue between studies is the non-standardized SES classification. Secondly, the lack of diversity in the study populations (for example sex) may mean that certain under-represented groups may have other unidentified factors that could influence outcomes. Moreover, factors relating to prolonged hospitalization as well as factors associated with PPM insertion were not reported in the individual studies. This gap in reporting may be further influenced by potential treatment biases based on SES. Patients from low SES background may present later, receive delayed care, or are managed with medical therapy rather than more aggressive interventions. Unconscious bias and resource availability can also contribute to these disparities, ultimately affecting both treatment decisions and outcomes.

## Conclusion

In conclusion, this meta-analysis underscores the profound socio-economic influences on TAVI outcomes, advocating for further research using standardised SES scoring and the development of specific interventions aimed at improving care for patients from less advantaged backgrounds. We have proposed a framework to better assess socio-economic risk with TAVI and would advocate that this SES data be collected in national registries and prospective research studies going forwards. This approach could both inform future research and potentially mitigate SES-related disparities in outcomes following TAVI.

## Abbreviations

AS: Aortic Stenosis
TAVI: Transcatheter Aortic Valve Implantation
SES: Socio-economic Status
MACE: Major Adverse Cardiovascular Events
PPM: Permanent Pacemaker Implantation
PRISMA: Preferred Reporting Items for Systematic Reviews and Meta-Analyses
MeSH: Medical Subject Headings
EMTREE: Embase Subject Headings
CVD: Cardiovascular Disease
HR: Hazard Ratio
RR: Risk Ratio
CI: Confidence Interval
ROBVIS: Risk of Bias Visualisation Tool
AVR: Aortic Valve Replacement
EU-CaRE: European Union – Cardiovascular Risk in the Elderly

## Acknowledgments

None.

## Data Availability Statement

The data underlying this article are available in the article and in its online supplementary material.

## Supplementary

**Supplementary Table S1:** Risk of bias assessment in Cohort studies.

|  | Selection population | Assessment of exposure | Blinding of outcome | Matching variables associated with outcome | Assessing prognostic factors | Outcome assessment | Follow up cohort adequate |
| --- | --- | --- | --- | --- | --- | --- | --- |
| 1. *Li et al 2024* | + | + | + | + | + | + | — |
| 2. *Hussain et al 2024* | + | + | ? | ? | + | + | — |
| 3. *Shawon et al 2024* | + | + | ? | ? | ? | ? | + |
| 4. *Brlecic et al 2023* | + | + | + | ? | + | + | + |
| 5. *Chabova et al 2022* | + | + | + | + | + | + | + |
| 6. *Bilfinger et al 2021* | + | + | + | ? | + | + | + |
| 7. *Kappelgaard et al 2021* | + | + | + | ? | ? | + | + |
| 8. *Farwati et al 2021* | + | + | + | + | + | + | ? |
| 9. *Mohee et al 2019* | + | + | + | + | + | + | + |
| 10. *Czarnecki et al 2017* | + | + | ? | ? | — | ? | ? |

Low risk of bias Unclear risk of bias High risk of bias

**Supplementary Table S2:** Characteristics of included studies.

| **Study;**<br>**(Country)** | **Study Design,**<br>**division of**<br>**SES** | **Size (n)\*\*** | **Period of**<br>**inclusion** | **Significant\***<br>**pre-procedural**<br>**characteristics**<br>**Low SES (n, %)** | **Significant\***<br>**pre-procedural**<br>**characteristics High**<br>**SES (n, %)** | **Primary**<br>**Outcome** | **Follow**<br>**up** |
| --- | --- | --- | --- | --- | --- | --- | --- |
| Li et al[16],<br>2024<br>(USA) | National,<br>retrospective,<br>division<br>according to<br>income and<br>zip code | Total: 27567<br>Low: 12355<br>High:15212 | 2015-2020 | Male N=6500 (52.61)<br>Caucasia N=9630 (77.94%)<br>DM N=2156 (17.45%)<br>Abuse. N=50, (0.4%)<br>HTN. N=8241(66.7%)<br>COPD N=3795 (30.72%)<br>H. BMI. N=2763 (22.36%)<br>PVD. N=2776 (22.47%)<br>CKD N=475, (3.84%)<br>Thyroid. N=2380 (19.26%)<br>CAD. N=8551 (69.21%)<br>pHTN. N=2084 (16.87%)<br>arrhyt. N=4447 (35.99%)<br>H.lipid. N=8801 (71.23%)<br>Prev MI N=1595, (12.91%)<br>Prev CABG N=2023, (16.37%) | 8660 (56.93)<br>13169 (86.57%)<br>2213 (14.55%)<br>39 (0.26%)<br>9617 (63.22%)<br>3602 (23.68%)<br>2617 (17.2%)<br>3253 (21.38%)<br>470 (3.09%)<br>3104 (20.4%)<br>10161 (66.8%)<br>2334 (15.34%)<br>5861 (38.53%)<br>11356 (74.65%)<br>1723 (11.33%)<br>2228(14.65%) | In hospital<br>mortality, MACE,<br>MI, CVA. | N/A |
| Hussain et<br>al[17], 2024<br>(USA) | National,<br>retrospective,<br>division<br>according to<br>income and<br>insurance | Total: 296740<br>Low: 62824<br>High: 77633 | 2016-2020 | N/A | N/A | Pacemaker<br>implantation | N/A |
| Shawon et<br>al[18], 2024<br>(Australia) | National,<br>retrospective,<br>division<br>according to<br>socio-economic<br>index for<br>areas | Total: 4198<br>Low: 641<br>High:1062 | 2013-2022 | N/A | N/A | 90 days and 1 year<br>mortality | 1 year |
| Brlecic et al[19],<br>2023<br>(USA) | National,<br>retrospective,<br>division<br>according to<br>pay,<br>education,<br>literacy,<br>housing,<br>employment<br>status | Total: 142786<br>Low: 27791<br>Medium:<br>80606<br>High: 34389 | 2016-2018 | Age 80 (74-76) years<br>Male N=14321, (51.5%)<br>CHF N=21723 (78.2%)<br>Arrhy N=15274, (55%)<br>COPD N=8957 (32.2%)<br>DM N=11478 (41.3%)<br>CKD N=10452, (37.6%)<br>H.BMI N=5975 (21.5%)<br>pHTN. N=5569 (20.0%)<br>H.lipid. N=19585 (70.5%)<br>Abuse. N=178, (0.6%) | 82 (76-87) years<br>N=19228, (55.9%)<br>N=24768 (72%)<br>N=20246, (58.9%)<br>N=8867 (25.8%)<br>N=11649 (33.9%)<br>N=11653, (33.9%)<br>N=5452 (15.9%)<br>N=6342 (18.4%)<br>N=25424 (73.9%)<br>N=123, (0.4%) | Procedural choice<br>SAVR vs. TAVI | 1 year |
| Chabova et<br>al[15], 2022<br>(Czech<br>Republic) | Single centre,<br>retrospective,<br>division<br>according to<br>education | Total: 439<br>Missing: 46<br>Low: 212<br>High: 167 | 2010-2020 | Age 77.4 ±7.2 years<br>Male N=88, (42%)<br>BMI 29.5 ±5.7<br>DM N=93, (43%)<br>HTN N=170 (80%)<br>PLT 209.1x109 /L<br>AR≥3 N=16 (8%) | 77.4 ±7.2 years<br>N=100, (60%)<br>28.3±4.8<br>N=51, (31%)<br>N=136 (81%)<br>188x109 /L<br>N=4 (2%) | all-cause mortality,<br>MACE | 2 years |
| Von<br>Kappelgaard et<br>al[12], 2021<br>(Denmark) | National,<br>retrospective,<br>division<br>according to<br>education | Total: 13861<br>Missing: 341<br>Low: 5782 | 2000-2016 | N/A | N/A | All-cause mortality | 1 year |
|  |  | Medium:<br>5238<br>High:<br>2500 |  |  |  |  |  |
| Bilfinger et al[13], 2021 (USA) | Institutional, prospective, division according to zip code via distressed community index (DCI) | Total: 286<br>DCI <10 (high): 73<br>DCI 10-20: 81<br>DCI 20-30:80<br>DCI>30 (low): 52 | Dec 2015- June 2018 | Female N=32 (61.5%)<br>CAD N=4 (7.7%)<br>AF N=12 (23.1%)<br>DM N=20 (38.5%) | N=36 (49.3%)<br>N=18 (24.7%)<br>N=33 (45.2%)<br>N=25 (34.2%) | All-cause mortality, MACE | 3 years |
| Farwati et al[11], 2021 (USA) | Nationwide, retrospective, division according to income | Total: 168853<br>Q1(low): 34835<br>Q2: 44370<br>Q3: 46007<br>Q4 (high): 43642 | Jan 2012- Dec 2017 | Age 81 (75-86)<br>Male N=18240 (52.4%)<br>DM N=13205 (37.9%)<br>HTN N= 29793 (85.5%)<br>AF N=11606 (33.3%)<br>BMI>30 N=5894 (16.9%) | 83 (77-88)<br>N=23516 (53.9%)<br>N=13956 (32%)<br>N= 36978 (84.7%)<br>N=16121 (36.9%)<br>N=5731 (13.1%) | MACE | 30 days |
| Mohee et al[10], 2019 (Wales) | Regional, retrospective, division according to zip code | Total: 387<br>Low: 91<br>High 296 | 2009-2018 | Age 80.4±9.3 years<br>Male N=46 (69.7%) | 83.5±7.9 years<br>153 (83.2%) | all-cause mortality, MACE | 3 years |
| Czarnecki et al[14], 2017 (Canada) | Regional, retrospective, division according to zip code | Total: 937<br>Unknown numbers Q1-Q5 | April 2007- March 2013 | - | - | Re-admission | 1 year |

\*Significant p<0.05
\*\*for the purpose of this study we have compared the Lowest SES with the Highest SES
Abbreviations; BMI=body mass index, DM=diabetes mellitus, HTN=hypertension, PLT=platelet, AR=aortic regurgitation, CV=cardiovascular, CAD=coronary artery disease, MS=marital status, AF=Atrial fibrillation, MACE=major adverse cardiac events, COPD=Chronic obstructive pulmonary disease, PVD=peripheral vascular disease, p.HTN=pulmonary hypertension, arrhyt=arrhythmia, h.lipid=hyperlipidaemia

